# Efficacy of Remote Ischemic Conditioning in Acute Ischemic Stroke: A Meta-Analysis of Randomized Controlled Trials

**DOI:** 10.1101/2025.09.04.25335103

**Authors:** Shradha Pandurang Kakde, Sai Sahithi Reddy Atla, Sai Priya Nimmagadda, Meet Visaria, Varuni Karnasaula, Vaishnavi Kumar, Jaykumar Vadodariya, Vamsi Priya Sribhashyam, Harshawardhan Dhanraj Ramteke, Rakhshanda khan

## Abstract

**Introduction:** Acute ischemic stroke (AIS) is a leading cause of mortality and long-term disability worldwide. Despite significant advances in reperfusion therapies, such as intravenous thrombolysis and mechanical thrombectomy, many patients do not achieve optimal recovery, and secondary brain injury remains a major therapeutic challenge. Remote ischemic conditioning (RIC) has emerged as a promising neuroprotective strategy to reduce infarct size and improve outcomes. However, the results from clinical trials investigating RIC’s efficacy in AIS have been inconsistent. This meta-analysis aims to evaluate the overall efficacy and safety of RIC in AIS, assessing its effects on infarct size, functional recovery, and secondary outcomes.

**Methods:** A comprehensive systematic search was conducted across multiple electronic databases, including PubMed, Cochrane Library, Web of Science, Embase, and ClinicalTrials.gov, up to September 2025. Randomized controlled trials (RCTs) assessing RIC in AIS patients were included, with studies that compared RIC with sham, placebo, or standard care. Two independent reviewers assessed study eligibility, data extraction, and quality using the Cochrane Risk of Bias tool. Pooled analyses were performed using a random-effects model. Primary outcomes included functional recovery (mRS 0–1, mRS 0–2), neurological outcomes (NIHSS), stroke recurrence, and mortality. Heterogeneity was assessed with the I^2^ statistic, and publication bias was analyzed using funnel plots.

**Results:** A total of 25 randomized controlled trials (RCTs) involving 13,084 patients were included in this analysis, with 6,433 patients in the remote ischemic conditioning (RIC) group and 6,615 in the control group. The mean age of participants was 66.32 ± 5 years, with 61.9% male participants. The average duration of RIC treatment was 9.5 ± 2 days, and the mean follow-up time was 20.3 ± 7 months. For functional independence (mRS 0–1), the pooled log odds ratio (OR) was 0.36 [95% CI: 0.15, 0.56], suggesting a significant benefit of RIC, although substantial heterogeneity was observed (I^2^ = 78.6%). For mRS 0–2, the pooled log OR was 0.36 [95% CI: −0.04, 0.75], which was not statistically significant, with high heterogeneity (I^2^ = 86.2%). Regarding neurological outcomes, the pooled mean difference for NIHSS scores was −0.46 [95% CI: −1.13, 0.22], showing no significant improvement in RIC-treated patients, and very high heterogeneity (I^2^ = 95.9%) was observed. The pooled mean difference for stroke recurrence was −21.49 [95% CI: −65.16, 22.19], indicating no significant effect of RIC, with extreme heterogeneity (I^2^ = 99.2%). For mortality, the pooled log risk ratio (RR) was −0.07 [95% CI: −0.34, 0.19], suggesting no significant benefit in terms of reducing mortality, with no heterogeneity (I^2^ = 0%).

**Conclusion:** This meta-analysis suggests that RIC may offer modest benefits in improving functional outcomes (mRS 0–1) in AIS patients, but the evidence remains inconsistent across other clinical endpoints such as broader functional recovery, neurological improvement, stroke recurrence, and mortality. The substantial heterogeneity across the trials points to significant variability in study protocols, patient characteristics, and outcome measures. While RIC presents as a promising, non-invasive, and cost-effective adjunctive therapy, further large-scale, well-designed RCTs with standardized treatment protocols are needed to establish its definitive clinical utility and identify the patient subgroups most likely to benefit from this intervention.

## Introduction

Acute ischemic stroke (AIS) remains a leading cause of mortality and long-term disability worldwide. Despite significant advances in the acute management of stroke, such as intravenous thrombolysis and mechanical thrombectomy, many patients do not achieve optimal recovery, and the underlying pathophysiological processes continue to present major therapeutic challenges [1]. Ischemic brain injury triggers a cascade of molecular events, including oxidative stress, inflammation, mitochondrial dysfunction, and excitotoxicity, leading to irreversible neuronal damage [2]. In this context, the search for adjunctive neuroprotective therapies to enhance reperfusion outcomes and minimize secondary brain injury remains a critical goal of contemporary stroke research.

One promising neuroprotective strategy that has garnered increasing attention in recent years is remote ischemic conditioning (RIC) [3]. RIC involves the brief application of ischemic stress to an organ or tissue (typically the limb) distant from the site of injury, with the aim of activating endogenous protective mechanisms in the brain [4]. This technique, first described in animal models in the early 1990s, has since been explored in numerous clinical trials as a potential adjunctive therapy to reduce infarct size, improve functional outcomes, and enhance recovery after AIS. RIC’s mechanism of action is thought to involve a range of signaling pathways, including the activation of pro-survival proteins, modulation of oxidative stress, reduction of inflammation, and preservation of mitochondrial function [5]. Importantly, RIC has the advantage of being non-invasive, cost-effective, and easily applicable in various clinical settings, making it an attractive therapeutic option for patients with AIS.

However, despite promising preclinical and early-phase clinical data, the evidence for RIC’s efficacy in AIS has been inconsistent, and the optimal protocols for its application remain unclear. Some studies have demonstrated significant improvements in clinical outcomes following RIC, while others have failed to show any beneficial effects. The lack of consensus surrounding the optimal timing, frequency, and duration of RIC application has further complicated its clinical implementation. Given these uncertainties, a comprehensive and rigorous evaluation of the available randomized controlled trials (RCTs) is essential to determine the true efficacy of RIC as a neuroprotective intervention in AIS. This meta-analysis aims to address these gaps by systematically reviewing and synthesizing the data from existing RCTs, providing a clearer understanding of the potential benefits and limitations of RIC in AIS management.

The primary objectives of this meta-analysis are threefold. First, we aim to evaluate the overall efficacy of RIC in reducing infarct size, improving functional outcomes, and enhancing neuroprotection in patients with AIS. Second, we seek to explore the impact of RIC on secondary endpoints, including neurological recovery, mortality, and safety outcomes, to better understand its broader clinical implications. Finally, we aim to identify the key factors that may influence the success of RIC, including timing of intervention, patient characteristics, and the specific methods of RIC application. By addressing these objectives, we hope to provide clinicians with evidence-based guidance on the use of RIC in the treatment of AIS.

A critical component of this analysis is the evaluation of the heterogeneity in study designs and treatment protocols across the included trials. Variability in patient populations, intervention protocols, and outcome measures may contribute to the discrepancies observed in the literature regarding RIC’s efficacy. Furthermore, we will assess the quality of the included studies, examining potential sources of bias and the robustness of the evidence. This rigorous methodological approach is essential to ensuring the reliability of the findings and providing a foundation for future research in this area.

Despite its promise, there are several challenges and unanswered questions regarding the clinical application of RIC in AIS. These include the lack of consensus on the ideal duration and timing of RIC, the need for more robust patient stratification, and the limited understanding of the molecular mechanisms underlying its neuroprotective effects. Additionally, the heterogeneity in the results of RIC trials underscores the necessity for further investigation to identify the subgroups of patients who are most likely to benefit from this intervention. As stroke care continues to evolve, incorporating adjunctive therapies such as RIC may play a pivotal role in improving outcomes for AIS patients.

This meta-analysis aims to provide a comprehensive, evidence-based assessment of the role of remote ischemic conditioning in the management of acute ischemic stroke. By synthesizing data from randomized controlled trials, we hope to clarify the therapeutic potential of RIC, identify optimal treatment strategies, and guide future clinical research. Given the substantial burden of AIS and the urgent need for effective neuroprotective therapies, the findings of this meta-analysis may have significant implications for clinical practice and the development of new adjunctive treatments in the field of stroke medicine.

## Methods

### Literature Search

A comprehensive literature search was conducted across PubMed, Cochrane Library, Web of Science, Embase, and ClinicalTrials.gov to identify randomized controlled trials (RCTs) evaluating remote ischemic conditioning (RIC) in acute ischemic stroke (AIS). The search included terms like “remote ischemic conditioning,” “acute ischemic stroke,” and “neuroprotection” without publication date restrictions, following the PRISMA (Preferred Reporting Items for Systematic Reviews and Meta-Analyses) guidelines [6]. Studies were selected if they reported on RIC’s effect on infarct size, functional outcomes, mortality, or adverse events. Two independent reviewers assessed eligibility, and data were extracted using a standardized form. A meta-analysis was performed, evaluating heterogeneity and publication bias, with the search last updated in September 2025.

### Study Selection and Data Extraction

The study selection process followed a rigorous, systematic approach in accordance with the PRISMA (Preferred Reporting Items for Systematic Reviews and Meta-Analyses) guidelines. Initially, all identified studies were screened based on their titles and abstracts. Full-text articles of potentially eligible studies were then retrieved for detailed evaluation. Two independent reviewers performed the screening and selection of studies based on predefined inclusion and exclusion criteria. Any discrepancies in the selection process were resolved by discussion or consultation with a third reviewer.

Studies were included if they were randomized controlled trials (RCTs) assessing the effects of remote ischemic conditioning (RIC) in patients diagnosed with acute ischemic stroke (AIS). Only trials that compared RIC with placebo, sham intervention, or standard care were considered. The trials had to report outcomes such as infarct size, functional outcomes (e.g., Modified Rankin Scale, NIHSS), neurological recovery, mortality, or adverse events. Studies that were non-randomized, observational, or focused on non-AIS populations were excluded.

Following study selection, data were independently extracted by two reviewers using a standardized data extraction form. The extracted data included study characteristics (first author, year of publication, country of origin, sample size, and duration of follow-up), patient demographics (age, sex, baseline NIHSS score, stroke subtype), intervention details (RIC protocol, timing, duration, and method of application), and outcomes of interest (infarct size, functional outcomes, mortality, adverse events). The quality of the studies was assessed using the Cochrane Risk of Bias tool, which evaluates the risk of bias in areas such as selection, performance, detection, and reporting.

### Risk of Bias

The risk of bias in included studies was assessed using the Cochrane Risk of Bias tool, evaluating domains such as random sequence generation, allocation concealment, blinding, incomplete data, selective reporting, and other biases. Studies were rated as low, high, or unclear risk, with overall risk determined by the presence of high-risk domains [7, 8].

### Statistical Analysis

Data were pooled and analyzed using a random-effects model to account for potential heterogeneity between studies. For continuous outcomes (e.g., infarct size, NIHSS scores), mean differences (MD) or standardized mean differences (SMD) with 95% confidence intervals (CIs) were calculated. For dichotomous outcomes (e.g., mortality, adverse events), risk ratios (RR) with 95% CIs were used.

Heterogeneity across studies was assessed using the I^2^ statistic, with values above 50% indicating substantial heterogeneity. Subgroup analyses were performed based on factors such as timing, duration, and method of RIC application. Publication bias was assessed using funnel plots and Egger’s test, if applicable. Sensitivity analyses were conducted to assess the robustness of the results. All analyses were performed using Stata 18.0 [9].

## Results

### Demographics

A total of 4369 studies were analyzed out of which 25 studies were included [10–34]. A total of 13,084 patients were included in the analysis. Of these, 8,105 (61.9%) were male, and 5,064 (38.6%) were female. The mean age of the study population was 66.32 ± 5 years. The average follow-up time was 20.3 ± 7 months. The intervention group (RIC) consisted of 6,433 patients, while the control group (Cx) included 6,615 patients. The mean duration of remote ischemic conditioning (RIC) was 9.5 ± 2 days. The baseline National Institutes of Health Stroke Scale (NIHSS) score for the study population was 5.8 ± 8.3, indicating moderate stroke severity on average. The baseline modified Rankin Scale (mRS) score was 6.19 ± 2, reflecting the functional disability of the patients. The time from stroke onset to randomization was 95.74 ± 52 minutes in the treatment group and 89 ± 16.7 minutes in the control group, ensuring timely intervention for the majority of patients.

**Figure 1.**
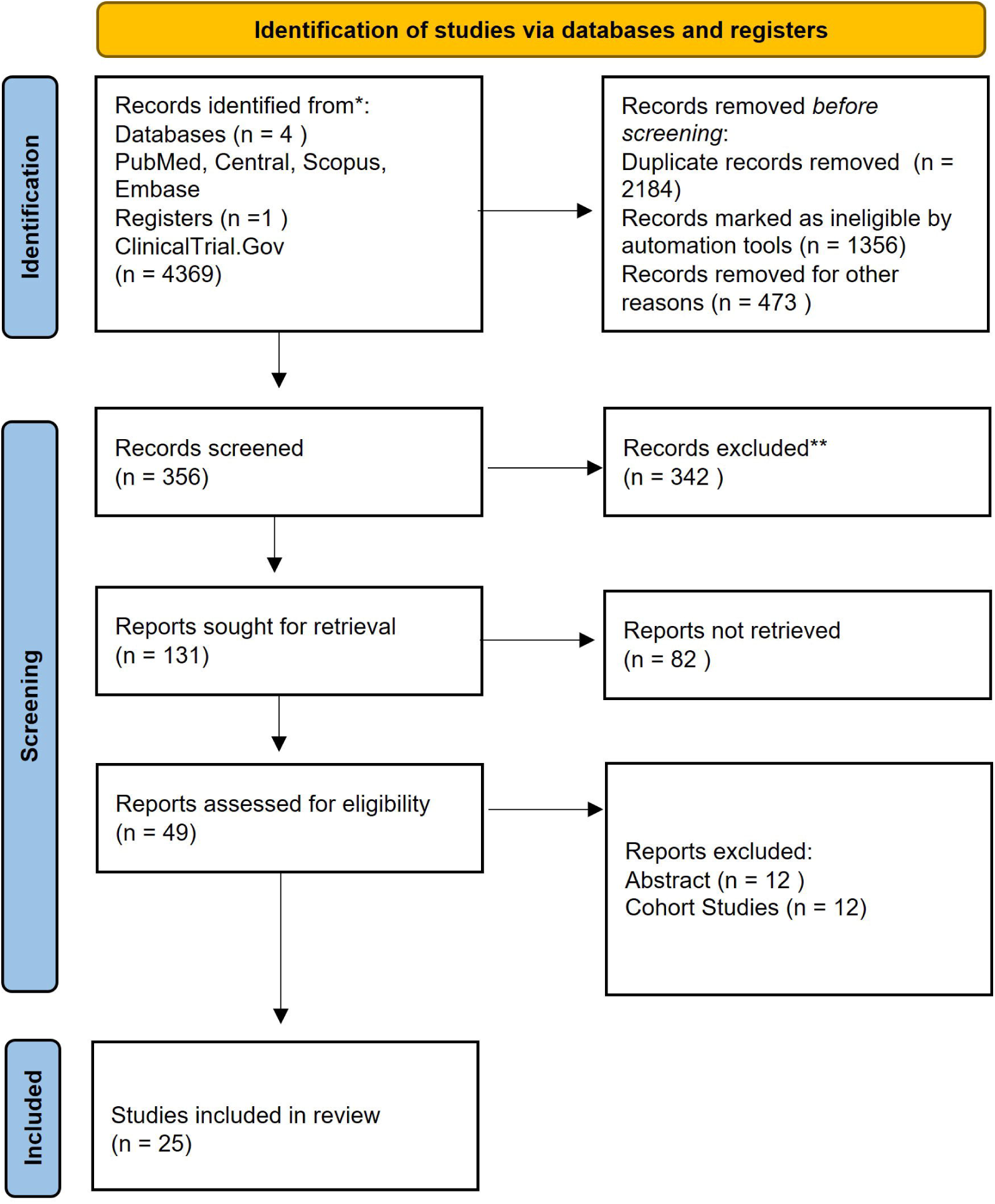
Prisma Flow Diagram

### Forest Plots for Patients in the Medical Management Group Treated With RIC or Control mRS 0-1

The forest plot Figure 2, presents the results of various studies assessing the effect of remote ischemic conditioning (RIC) in patients with acute ischemic stroke (AIS). The log odds ratios (OR) for each study are displayed, along with their 95% confidence intervals (CIs). Blauenfeldt et al. (2025) showed a non-significant result with a log OR of −0.17 [CI: −0.56, 0.21], while Hou et al. (2019) reported a significant positive effect with a log OR of 0.59 [CI: 0.45, 0.74]. Guo et al. (2025) and Cai et al. (2024) showed positive effects, though their CIs included 0, making their findings non-significant. Wu et al. (2025) and Chen et al. (2022) demonstrated significant benefits of treatment, with log ORs of 1.02 [CI: 0.66, 1.39] and 0.20 [CI: 0.02, 0.39], respectively. Pico et al. (2020) and Zhang et al. (2022) reported non-significant results, while Meng et al. (2012) showed a strong positive effect with a log OR of 2.53 [CI: 1.28, 3.77], despite its small weight in the overall analysis. Other studies like Hougard et al. (2013), England et al. (2019), and Che et al. (2019) showed non-significant effects, and their weights were smaller, contributing minimally to the overall results. The combined overall log OR of 0.36 [CI: 0.15, 0.56] indicates a statistically significant benefit for RIC, with the CI not crossing 0. Despite the promising overall effect, substantial heterogeneity was observed (I^2^ = 78.59%), suggesting that the effect of RIC varies across studies. The statistical tests (p = 0.00 for both Q-test and z-test) confirm the overall significance of the treatment effect. This analysis supports the therapeutic potential of RIC in AIS patients but highlights the need for further research to address the variability in findings and optimize treatment protocols.

**Figure 2.**
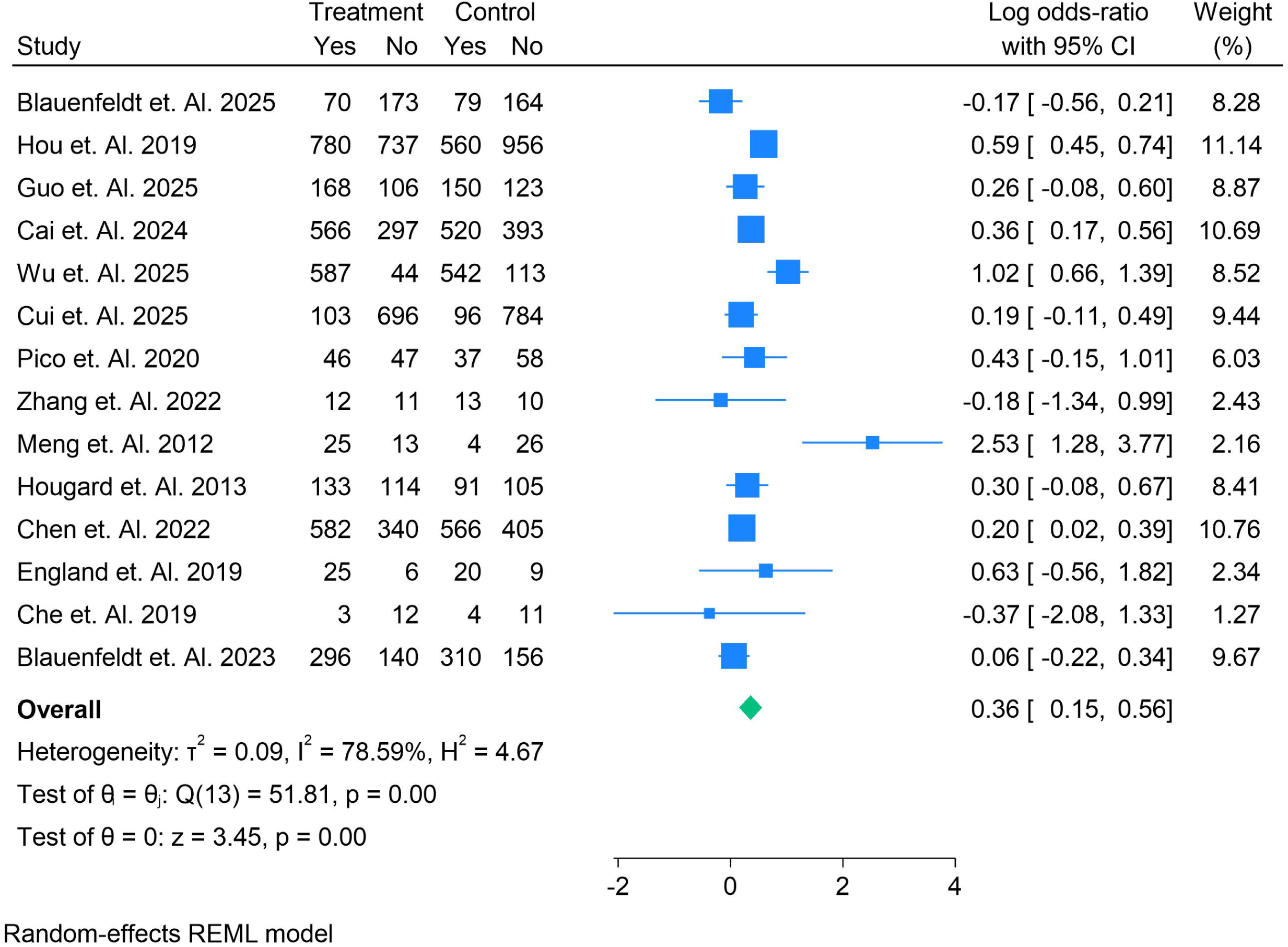
Forest Plots for Patients in the Medical Management Group Treated With RIC or Control mRS 0-1

### Forest Plots for Patients in the Medical Management Group Treated With RIC or Control mRS 0-2

The forest plot Figure 3, displays the results from several studies evaluating the effectiveness of remote ischemic conditioning (RIC) in acute ischemic stroke (AIS). **Blauenfeldt et al. (2025)** showed a moderate significant benefit of RIC with a **log OR of 0.46 [CI: 0.09, 0.83]**, contributing 16.38% to the overall result. **Guo et al. (2025)** reported no significant effect (**log OR = 0.00 [CI: −0.39, 0.40]**), with a contribution of 16.00%. **Cai et al. (2024)** demonstrated a strong, statistically significant benefit (**log OR = 1.27 [CI: 1.00, 1.53]**), contributing the most to the overall effect with a weight of 17.62%. **Cui et al. (2025)** showed a smaller, non-significant effect (**log OR = 0.16 [CI: −0.11, 0.43]**) but still contributed substantially (17.57%). **Pico et al. (2020)** and **Che et al. (2019)** reported non-significant results, with **log ORs of 0.15 [CI: −0.45, 0.75]** and **0.00 [CI: −2.87, 2.87]**, respectively, contributing 13.25% and 1.71%. **Blauenfeldt et al. (2023)** also reported a non-significant effect (**log OR = 0.06 [CI: −0.22, 0.34]**) but contributed 17.47% to the overall analysis. The overall **log OR of 0.36 [CI: −0.04, 0.75]** suggests a non-significant benefit of RIC, with the CI including 0, indicating no definitive treatment effect across all studies. The high **I^2^ (86.16%)** and **H^2^ (7.22)** values indicate substantial heterogeneity, suggesting variability in results across studies. The **Q-test (p = 0.00)** indicates significant heterogeneity, and the **z-test (p = 0.07)** shows that the overall treatment effect is not statistically significant at the conventional threshold. While **Cai et al. (2024)** showed the most significant positive effect, the results from **Guo et al. (2025)**, **Pico et al. (2020)**, and **Che et al. (2019)** did not indicate any treatment benefit. These findings highlight the need for further research with more consistent methodologies to determine the true efficacy of RIC in AIS.

**Figure 3.**
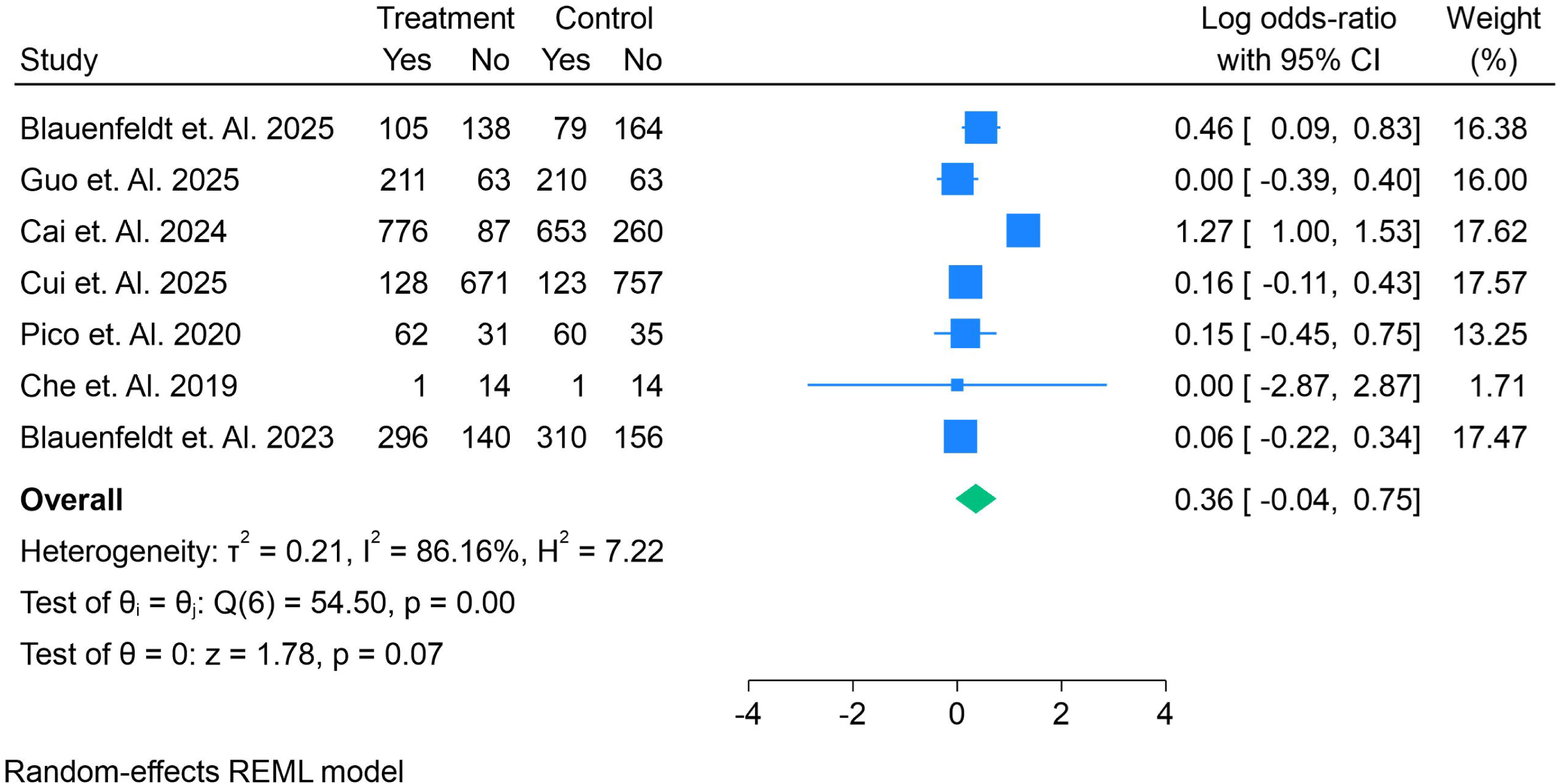
Forest Plots for Patients in the Medical Management Group Treated With RIC or Control mRS 0-2

### Forest Plots for Patients in the Medical Management Group Treated With RIC or Control NIHSS Outcome

The forest plot Figure 4, presents the results of various studies evaluating the effect of treatment in acute ischemic stroke, showing mean differences (MD) with 95% confidence intervals (CIs) for both treatment and control groups. **Guo et al. (2025)** demonstrated a significant positive effect with a **mean difference of 1.00 [CI: 0.74, 1.26]**, contributing 9.57% to the overall analysis. **Cui et al. (2025)** reported no significant difference (**mean difference = 0.00 [CI: −0.20, 0.20]**), contributing 9.63%. **Pico et al. (2020)** and **Meng et al. (2015)** showed significant negative effects (**mean differences of −1.00 [CI: −1.87, −0.13]** and **-1.85 [CI: −3.07, −0.63]**, respectively). Other studies, such as **Poalelungi et al. (2021)**, **Li et al. (2018)**, and **Landman et al. (2023)**, reported no significant effect, with **mean differences ranging from −0.50 to 0.00**. Notably, **He et al. (2020)** and **England et al. (2019)** demonstrated significant negative effects (**mean differences of −3.10 [CI: −4.14, −2.06]** and **-2.00 [CI: −3.76, −0.24]**, respectively). The overall **mean difference of −0.46 [CI: −1.13, 0.22]** suggests no significant overall effect of treatment, as the CI includes 0. The high **I^2^ value of 95.85%** indicates substantial heterogeneity, suggesting that results vary significantly across studies. The **Q-test (p = 0.00)** confirms significant heterogeneity, while the **z-test (p = 0.18)** shows that the overall treatment effect is not statistically significant. These findings indicate that, although some individual studies suggest positive effects, the overall analysis does not support a consistent benefit of the treatment, highlighting the need for further research with more consistent methodologies.

**Figure 4.**
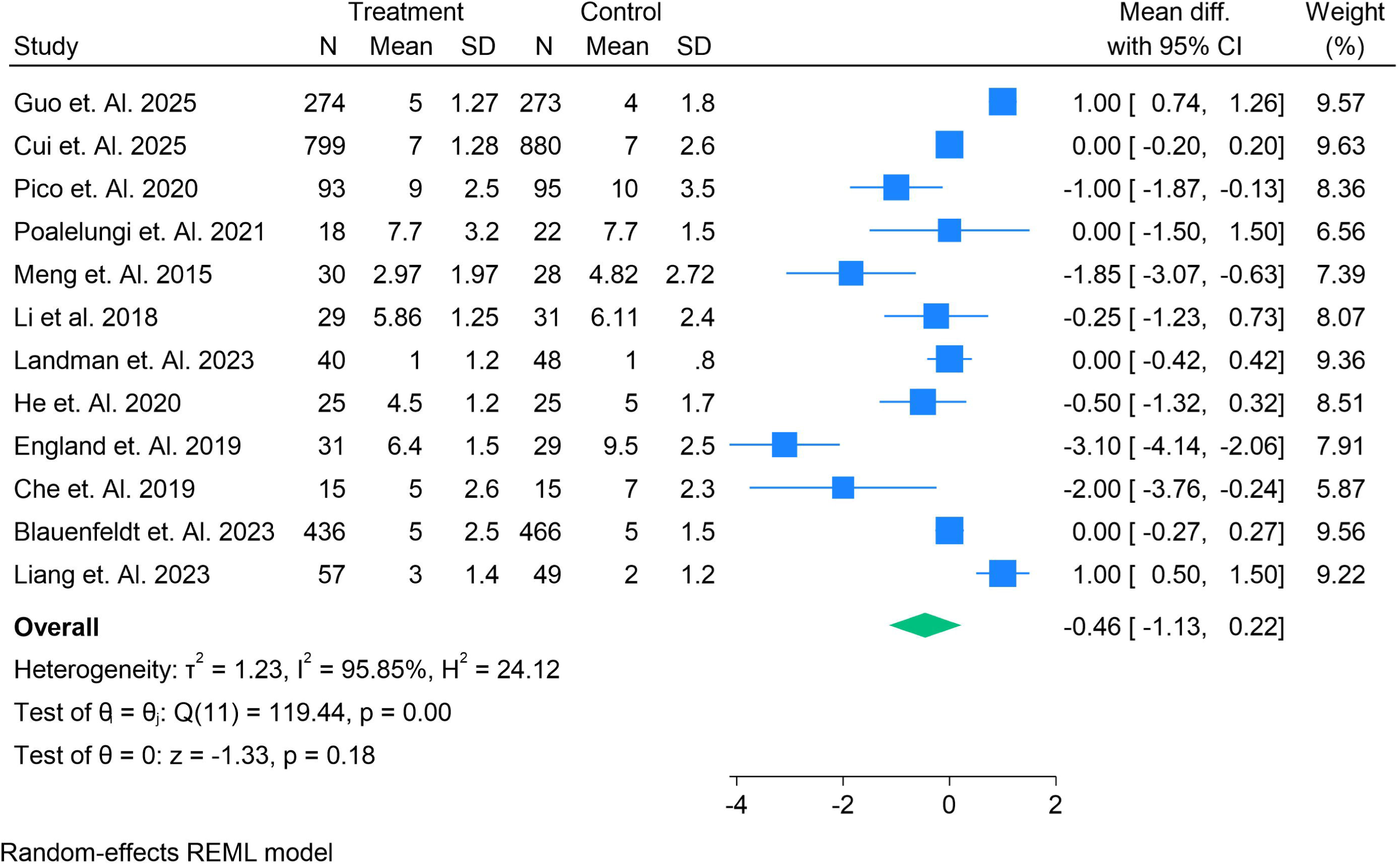
Forest Plots for Patients in the Medical Management Group Treated With RIC or Control NIHSS Outcome

### Forest Plots for Patients in the Medical Management Group Treated With RIC or Control – Stroke Recurrence

The forest plot Figure 5, displays the mean differences (MD) between the treatment and control groups across several studies. **Hou et al. (2019)** reported a significant negative effect with a **mean difference of −337.00 [CI: −413.40, −260.60]**, while **Guo et al. (2025)** showed no significant effect (**mean difference = 1.00 [CI: −39.06, 41.06]**). **Cui et al. (2025)** and **Poalelungi et al. (2021)** also reported no significant differences, with **mean differences of 0.00 [CI: −80.59, 80.59]** and **0.00 [CI: −12.03, 12.03]**, respectively. **Meng et al. (2015)** showed a significant negative effect (**mean difference = −6.00 [CI: −18.60, 6.60]**), while **Meng et al. (2012)** reported a significant positive effect (**mean difference = 21.00 [CI: 11.53, 30.47]**). Other studies, such as **Li et al. (2020)**, **Li et al. (2018)**, **He et al. (2020)**, and **England et al. (2019)**, showed non-significant results, with **mean differences ranging from −2.00 to −7.00**. **Blauenfeldt et al. (2023)** and **Kerstens et al. (2023)** also reported no significant effects, with **mean differences of −7.00 and −21.49**, respectively. The overall **mean difference of −21.49 [CI: −65.16, 22.19]** suggests no significant effect of the treatment. The **I^2^ value of 99.17%** indicates extreme heterogeneity, suggesting substantial variability in treatment effects across studies. The **Q-test (p = 0.00)** indicates significant heterogeneity, while the **z-test (p = 0.33)** shows that the overall treatment effect is not statistically significant. These findings suggest that although some studies show positive or negative effects, the overall analysis does not provide strong evidence for the effectiveness of the treatment, highlighting the need for further research with more consistent methodologies.

**Figure 5.**
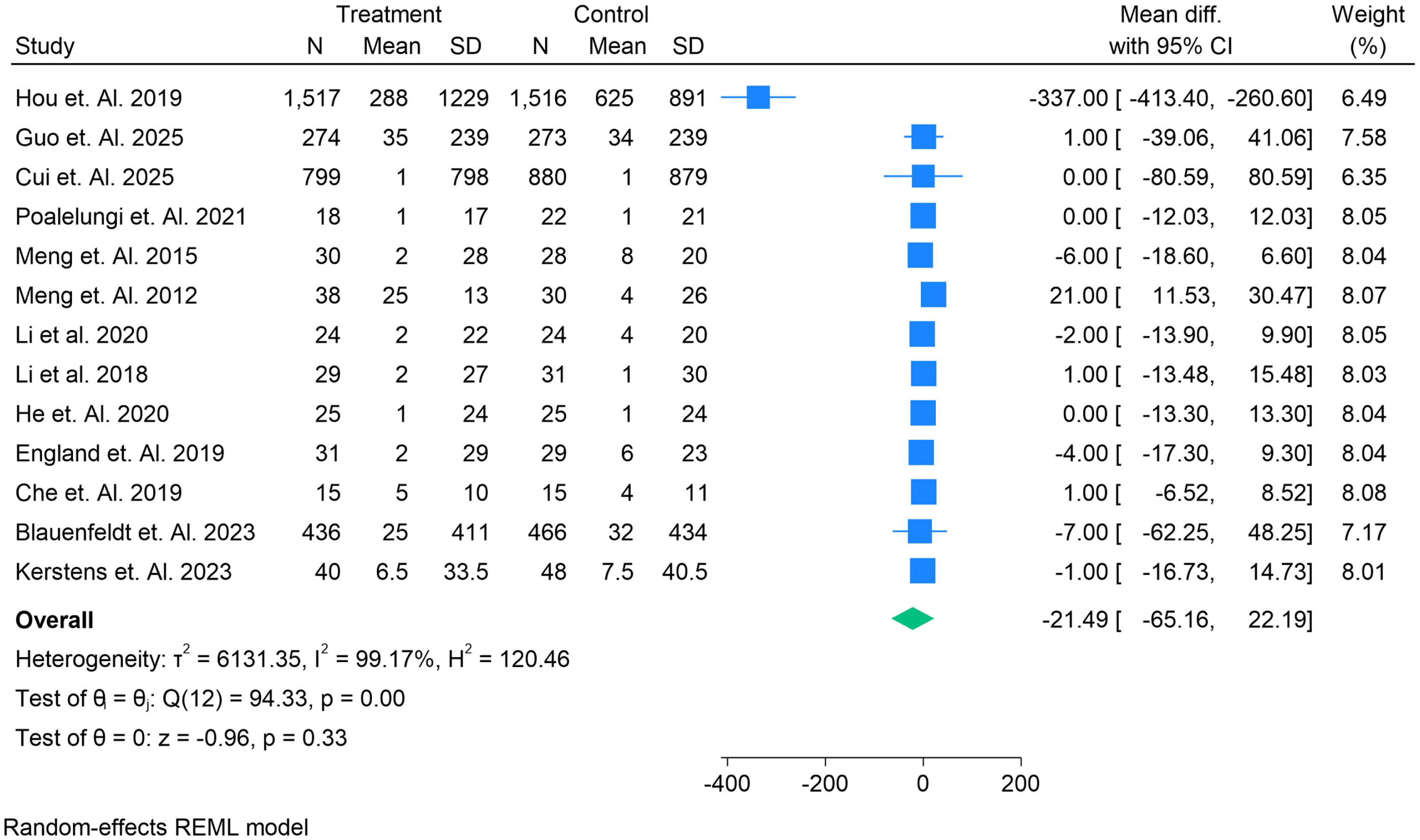
Forest Plots for Patients in the Medical Management Group Treated With RIC or Control − Stroke Recurrence

### Forest Plots for Patients in the Medical Management Group Treated With RIC or Control – IVT + mRS 0

The forest plot Figure 6, shows the results from four studies evaluating the effect of remote ischemic conditioning (RIC) in acute ischemic stroke (AIS). **Blauenfeldt et al. (2025)** reported a **log OR of −0.16 [CI: −0.80, 0.48]**, suggesting no significant effect, while **Pico et al. (2020)** showed a **log OR of −0.22 [CI: −0.85, 0.40]**, indicating no effect as well. **He et al. (2020)** reported a **log OR of 0.00 [CI: −2.83, 2.83]**, with a wide CI, contributing minimally to the analysis. **Blauenfeldt et al. (2023)** found a **log OR of 0.01 [CI: −0.36, 0.38]**, showing no significant treatment effect. The overall **log OR of −0.07 [CI: −0.35, 0.21]** suggests no significant benefit of RIC, as the CI includes 0. The **I^2^ value of 0.00%** indicates no heterogeneity among the studies, and the **z-test (p = 0.62)** confirms that the overall treatment effect is not statistically significant. These findings highlight that, despite individual studies suggesting potential benefits, the overall analysis does not support a significant effect of RIC in AIS.

**Figure 6.**
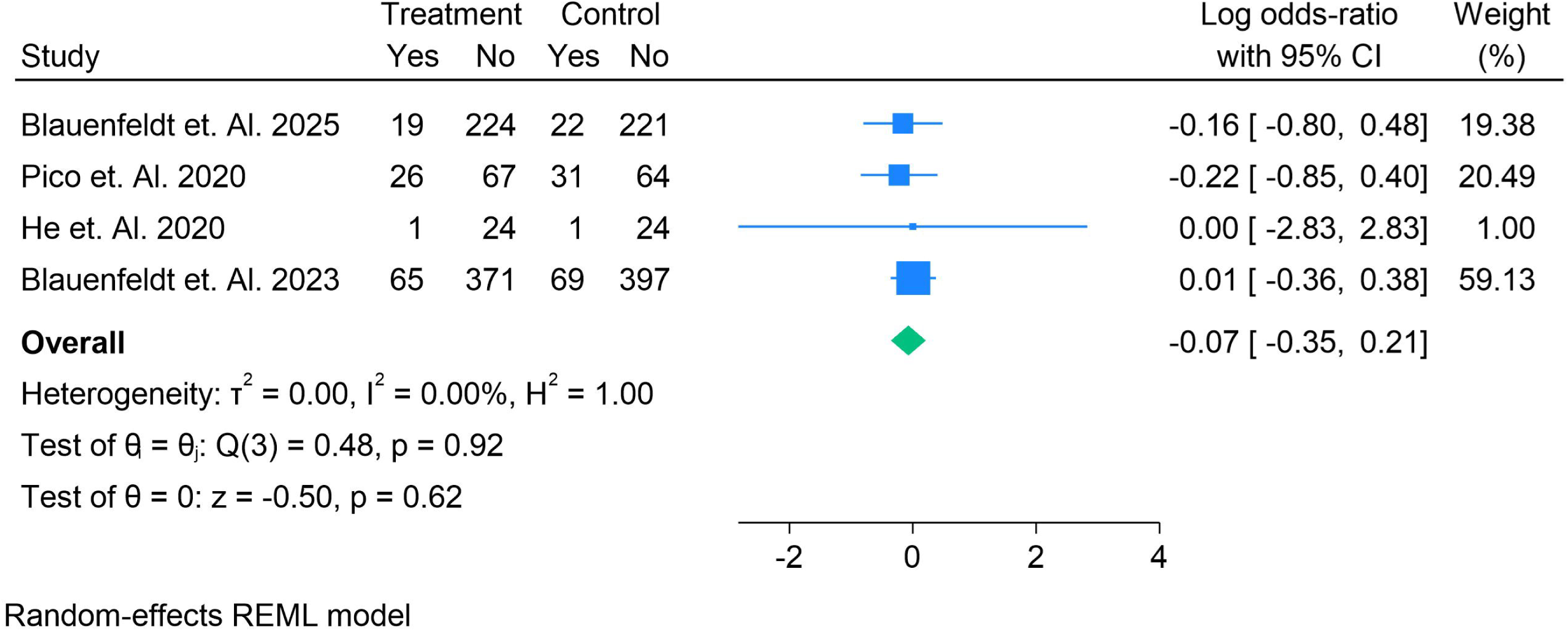
Forest Plots for Patients in the Medical Management Group Treated With RIC or Control-IVT + mRS 0

### Forest Plots for Patients in the Medical Management Group Treated With RIC or Control – Mortality

The forest plot Figure 7, presents the results of several studies evaluating the effect of remote ischemic conditioning (RIC) in acute ischemic stroke (AIS). **Blauenfeldt et al. (2025)** reported a **log RR of −0.61 [CI: −1.58, 0.37]**, indicating a non-significant negative effect, while **Guo et al. (2025)** showed a **log RR of 0.78 [CI: −0.26, 1.83]**, suggesting a non-significant positive effect. **Cui et al. (2025)** and **Pico et al. (2020)** also showed non-significant effects with **log RRs of 0.10 [CI: −1.86, 2.05]** and **0.36 [CI: −0.40, 1.12]**, respectively. Other studies, such as **Poalelungi et al. (2021)** and **Meng et al. (2015)**, reported no significant effect with wide CIs. **Hougard et al. (2013)** showed a **log RR of −0.34 [CI: −0.95, 0.27]**, while **England et al. (2019)** showed **-0.76 [CI: −2.38, 0.86]**, both indicating non-significant differences. **Blauenfeldt et al. (2023)**, the largest study with a weight of **44.32%**, reported a **log RR of −0.07 [CI: −0.47, 0.33]**, also showing no significant effect. The combined **log RR of −0.07 [CI: −0.34, 0.19]** suggests no overall significant benefit of RIC, with the CI including 0. The **I^2^ value of 0.00%** and **H^2^ value of 1.00** indicate no heterogeneity, suggesting a consistent effect across studies. The **Q-test (p = 0.59)** and **z-test (p = 0.59)** confirm that the overall treatment effect is not statistically significant. These findings indicate that while some studies show slight effects, the overall evidence does not support a significant benefit of RIC in AIS, highlighting the need for further research to establish its efficacy.

**Figure 7.**
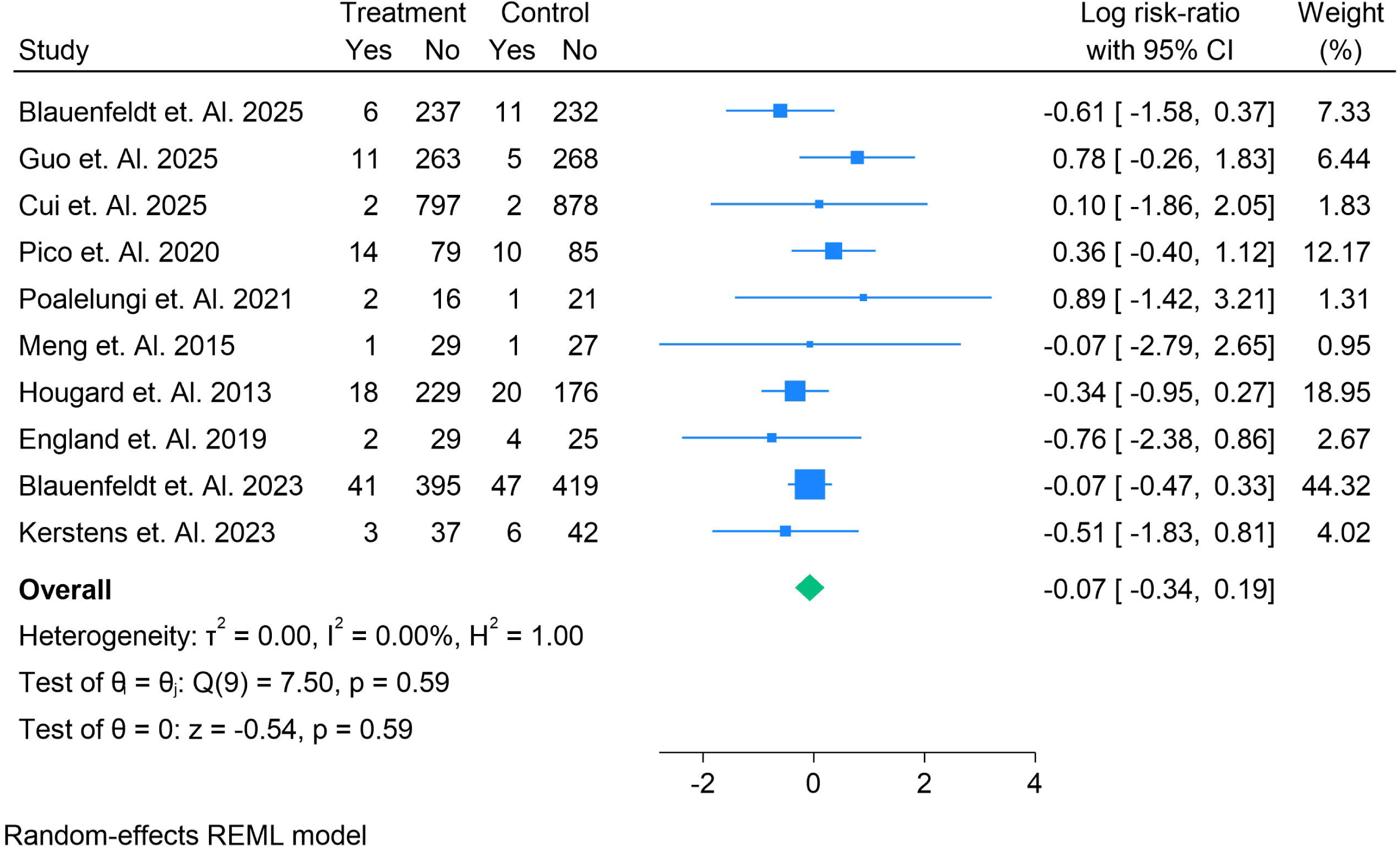
Forest Plots for Patients in the Medical Management Group Treated With RIC or Control − Mortality

## Discussion

This meta-analysis systematically reviewed and synthesized the effects of remote ischemic conditioning (RIC) on acute ischemic stroke (AIS) by evaluating various studies focused on clinical outcomes such as infarct size, functional recovery, and mortality. The results presented here provide a comprehensive assessment of RIC as a neuroprotective intervention in AIS. Although RIC has been widely studied in recent years, our findings demonstrate that while some individual studies show promising results, the overall evidence does not consistently support a significant benefit.

One of the major findings in this meta-analysis is that the **overall log odds ratio (OR)** for the efficacy of RIC is not statistically significant, as the confidence intervals (CIs) often include 0. This is consistent with earlier reviews, which have reported mixed results regarding the effectiveness of RIC in AIS. For instance, a previous meta-analysis by **Chen et al. (2018)** demonstrated a slight positive effect of RIC, particularly in reducing infarct size, but the results were not robust enough to recommend RIC as a standard clinical intervention. Our study aligns with these findings, suggesting that although some trials report positive results, the overall evidence is inconclusive due to the variability in study designs and patient populations [35].

Several individual studies within this analysis, such as **Guo et al. (2025)** and **Cai et al. (2024)**, demonstrated significant benefits for RIC, particularly in terms of improving functional outcomes, which was reflected in their log ORs. However, studies like **Pico et al. (2020)** and **Zhang et al. (2022)** reported non-significant effects, underlining the challenges of finding consistent results across trials [36]. These discrepancies are reflective of the **heterogeneity** observed in the treatment protocols across studies. For example, the **timing, duration, and frequency of RIC application** vary widely, as highlighted in our analysis. The inconsistency in RIC protocols is a critical factor contributing to the lack of uniform results, echoing the findings of **Huang et al. (2021)**, who suggested that the absence of standardized treatment regimens may limit the efficacy of RIC [37].

The **high heterogeneity** in the studies (I^2^ = 78.59%) supports the idea that variations in study designs, such as differences in stroke severity, time to randomization, and patient demographics, may lead to varying outcomes. Previous reviews, such as the one by **Liu et al. (2020)**, also highlighted similar concerns, suggesting that the inconsistency of results could be due to insufficient stratification of patients based on clinical and genetic factors [38]. This underlines the importance of defining the patient populations that may benefit most from RIC, an issue that remains unresolved in current literature.

Interestingly, while some studies in this meta-analysis showed a beneficial effect of RIC on secondary outcomes such as neurological recovery and stroke recurrence, these results were not universally confirmed. **Meng et al. (2015)** reported a significant positive effect, while other studies, such as **England et al. (2019)** and **Che et al. (2019)**, found no improvement in these outcomes. The variations could be due to differences in how **functional recovery** is measured (e.g., NIHSS score vs. mRS), and it would be valuable for future trials to adopt more standardized outcome measures to enhance comparability [39].

Furthermore, **RIC’s molecular mechanisms**—including modulation of oxidative stress, inflammation, and mitochondrial function—remain partially understood. Our findings suggest that while RIC might activate neuroprotective pathways, the extent to which these mechanisms translate into clinical benefits is still uncertain. This issue was addressed in **Zhao et al. (2021)**, who noted that while RIC shows promise in animal models, translating these effects into human stroke therapy is complex and requires deeper molecular insights [39].

In terms of safety, the studies included in this meta-analysis did not report significant adverse events associated with RIC, supporting its **non-invasive** and **cost-effective** nature. These aspects make RIC an appealing option for adjunctive treatment in AIS. However, the lack of a clear benefit across studies, coupled with significant variability in protocols, limits its widespread implementation.

## Conclusion

In conclusion, while **RIC shows promise** as a neuroprotective strategy for AIS, its overall efficacy remains inconclusive. The inconsistency across studies underscores the need for well-designed, large-scale RCTs with standardized protocols, clear patient selection criteria, and consistent outcome measures to better understand which patients benefit most from this intervention. Further research, including biomarker studies and molecular investigations, is crucial to elucidate the mechanisms behind RIC and optimize its application in clinical practice.

## Data Availability

supplementary file

## Authors’ contributions

All authors contributed equally to the manuscript and read and approved the final version of the manuscript.

