## supplementary file for "Efficacy of Remote Ischemic Conditioning in Acute Ischemic Stroke: A Meta-Analysis of Randomized Controlled Trials"

Supplementary Files

Search String

(((((((((((((((((((((((((((((Strokes[Title/Abstract]) OR (Cerebrovascular Accident[Title/Abstract])) OR (Cerebrovascular Accidents[Title/Abstract])) OR (CVAs (Cerebrovascular Accident[Title/Abstract]))) OR (Cerebrovascular Apoplexy[Title/Abstract])) OR (Apoplexy, Cerebrovascular[Title/Abstract])) OR (Vascular Accident, Brain[Title/Abstract])) OR (Brain Vascular Accident[Title/Abstract])) OR (Brain Vascular Accidents[Title/Abstract])) OR (Vascular Accidents, Brain[Title/Abstract])) OR (Cerebrovascular Stroke[Title/Abstract])) OR (Cerebrovascular Strokes[Title/Abstract])) OR (Stroke, Cerebrovascular[Title/Abstract])) OR (Strokes, Cerebrovascular[Title/Abstract])) OR (Apoplexy[Title/Abstract])) OR (Cerebral Stroke[Title/Abstract])) OR (Cerebral Strokes[Title/Abstract])) OR (Stroke, Cerebral[Title/Abstract])) OR (Strokes, Cerebral[Title/Abstract])) OR (Stroke, Acute[Title/Abstract])) OR (Acute Stroke[Title/Abstract])) OR (Acute Strokes[Title/Abstract])) OR (Strokes, Acute[Title/Abstract])) OR (Cerebrovascular Accident, Acute[Title/Abstract])) OR (Acute Cerebrovascular Accident[Title/Abstract])) OR (Acute Cerebrovascular Accidents[Title/Abstract])) OR (Cerebrovascular Accidents, Acute[Title/Abstract])) OR ("Stroke"[Mesh]))) AND (((((((((remote ischemic conditioning [Title/Abstract]) OR (RIC[Title/Abstract])) OR (remote ischemic preconditioning [Title/Abstract])) OR (RIPreC[Title/Abstract])) OR (remote ischemic perconditioning[Title/Abstract])) OR (RIPerC[Title/Abstract])) OR (remote ischemic postconditioning[Title/Abstract])) OR (RIPostC[Title/Abstract])))

Table S1. Summary of findings and baseline characteristics

| **Sr no** | **Author Name year** | **Country of Trial** | **Total Sample** | **Male** | **Female** | **Mean Age** | **Follow Up Time** | **Rx Name** | **Rx Sample** | **Cx Name** | **Cx Sample** | **Type of Disease** | **RIC Duration** | **NIHSS Baseline Score** | **mRS base line score** | **Stroke to Randomisation Time min** | **Stroke to Randomisation Time min** | **RIC Treatment Methods** | **GRADE** | **Main finding** |
| --- | --- | --- | --- | --- | --- | --- | --- | --- | --- | --- | --- | --- | --- | --- | --- | --- | --- | --- | --- | --- |
| 10 | Blauenfeldt et. Al. 2025 | Denmark | 486 | 279 | 207 | 72 | 24 | RIC | 243 | Sham | 243 | AIS | 7 days | 5 | 5 | 62 | 53 | **Bilateral upper limbs**, **5 cycles**, **50 minutes**, **2 times per day**. | Moderate | RIC treatment significantly reduced erythrocyte aggregation rate but had no significant impact on RBC deformability, NO content, or nitrite levels in patients with AIS. |
| 11 | Hou et. Al. 2019 | China | 3033 | 1954 | 1079 | 61.1 | 42 | RIC | 1517 | Sham | 1516 | ICAS | 12 | 1 | 1 | 12 | 12 | Bilateral upper limbs, 5 cycles, 50 min, 1 time per day for 12 months . | High | RIC significantly reduced the recurrence of ischemic stroke in patients with symptomatic ICAS, especially for those who complied with the therapy . |
| 12 | Guo et. Al. 2025 | China | 558 | 379 | 168 | 64 | 3 | RIC | 274 | Sham | 273 | AIS | 7 | 6 | 6 | 810 | 810 | Unilateral upper limb, cuff pressure 200 mm Hg, twice daily for 7 days, 5 minutes ischemia followed by 5 minutes reperfusion | High | Remote ischemic conditioning (RIC) was safe in patients with acute ischemic stroke treated with intravenous thrombolysis (IVT), but it did not significantly improve functional outcomes |
| 13 | Cai et. Al. 2024 | China | 1776 | 1170 | 606 | 65 | 3 | RIC | 863 | Sham | 913 | AIS | 14 | 6 | 6 | 48 | 48 | **5 minutes, totaling 50 minutes**, administered twice daily for 10–14 days | High | RIC improves functional outcomes, especially in patients without hypertension history, with a higher proportion achieving an mRS of 0–1 at 90 days . |
| 14 | Wu et. Al. 2025 | China | 1286 | 837 | 449 | 60.8 | 3.3 | RIC | 631 | Sham | 655 | TIA | 12 |  |  | 80 | 80 | 5 cycles per session, Each cycle: 5 minutes of inflation at 200 mmHg followed by 5 minutes of deflation, Once daily treatment for a year | High | Elevated lipoprotein(a) levels independently predicted recurrent ischemic stroke risk, and remote ischemic conditioning reduced stroke recurrence in patients with higher lipoprotein(a) levels. |
| 15 | Cui et. Al. 2025 | China | 1679 | 839 | 840 | 65 | 90 | RIC | 799 | Sham | 880 | AIS | 14 | 7 | 7 | 25.7 | 25.7 | 5 cycles, 200 mm Hg for 5 minutes, 5 minutes deflation, twice daily for 10–14 days | High | Remote Ischemic Conditioning (RIC) treatment improved 90-day functional outcomes more significantly in patients with low platelet-to-neutrophil ratios (PNR |
| 16 | Moyle et. Al. 2023 | United Kingdom | 24 | 16 | 8 | 58.6 | 2.1 | RIC | 12 | Sham | 12 | PSF | 6 | 5 | 5 | 83 | 83 | Blood pressure cuff inflated to 200 mmHg for 5 min, deflation for 5 min, repeated 4 cycles, 3 times per week for 6 weeks. | High | RIC is safe, acceptable, and feasible for people with post-stroke fatigue and may reduce fatigue severity. |
| 17 | Pico et. Al. 2020 | France | 188 | 98 | 90 | 67.2 | 3 | RIC | 93 | Sham | 95 | AIS | 3.7 | 9 | 10 | 105 | 103 | 4 cycles of 5-minute inflations and 5-minute deflations to the thigh at 110 mm Hg above systolic BP | High | Remote ischemic perconditioning did not reduce brain infarction growth at 24 hours after symptom onset |
| 18 | Zhang et. Al. 2022 | China | 46 | 26 | 15 | 63.1 | 90 | RIC | 23 | Sham | 23 | AIS | 6 | 6 | 6 | 48 | 48 | Unilateral upper arm, 5 cycles, inflation to 200 mmHg, deflation for 5 min, twice daily for 6 consecutive days. | Moderate | Remote Ischemic Conditioning (RIC) is safe in the prevention of Stroke-Associated Pneumonia (SAP) in patients with acute ischemic stroke. |
| 19 | Poalelungi et. Al. 2021 | Romania | 40 | 24 | 16 | 65 | 180 | RIC | 18 | Sham | 22 | AIS | 5 | 7.7 | 7.7 | 24 | 24 | 5 cycles of cuff inflation for 3 minutes, followed by 5 minutes of reperfusion, twice daily for the first 5 days | Moderate | Remote ischemic conditioning may improve disability and cognition in acute ischemic stroke, though differences did not reach statistical significance |
| 20 | Meng et. Al. 2015 | China | 58 | 38 | 20 | 83.5 | 6 | RIC | 30 | Sham | 28 | SIAS | 4.5 | 4 | 6 | 4.7 | 4.7 | 5 cycles of bilateral upper arm ischemia followed by 5 minutes of reperfusion, twice daily for 180 days. | High | BAIPC may safely reduce stroke recurrence in octo- and nonagenarians with symptomatic IAS. |
| 21 | Meng et. Al. 2012 | China | 68 | 40 | 28 | 60 | 10 | RIC | 38 | Sham | 30 | TIA | 16 | 7 | 8 | 10.8 | 11.3 | 5 cycles of bilateral upper limb ischemia for 5 minutes, followed by 5 minutes of reperfusion, performed twice a day for 300 days | Moderate | Bilateral arm ischemic preconditioning (BAIPC) significantly reduced stroke recurrence and improved cerebral perfusion in patients with intracranial arterial stenosis |
| 22 | Li et al. 2020 | China | 48 | 30 | 18 | 67.5 | 6 | RIC | 24 | Sham | 24 | AIS | 3 | 7.13 | 7.58 | 183 | 183 | 4 cycles of intermittent limb ischemia; 5 min inflation (20–30 mmHg above systolic blood pressure) and 5 min deflation | Moderate | Remote Ischemic Post-Conditioning (RIPC) was well tolerated and may improve cognitive and neurological outcomes in patients with post-stroke cognitive impairment |
| 23 | Li et al. 2018 | China | 60 | 29 | 31 | 65 | 3 | RIC | 29 | Sham | 31 | AIS | 14 | 5.86 | 7 | 39.7 | 42.16 | 4 cycles of intermittent limb ischemia, alternating 5 minutes inflation (20 mm Hg above systolic blood pressure) and 5 minutes deflation | Moderate | LIPostC after acute stroke is well tolerated, appears safe, and may improve neurological outcomes and decrease cerebral infarct volume |
| 24 | Landman et. Al. 2023 | Netherlands | 88 | 60 | 28 | 72.3 | 12 | RIC | 40 | Sham | 48 | AIS | 1 | 5 | 6.5 | 118.8 | 14.1 | Four cycles of 5-min inflation of the blood pressure cuff at 200 mm Hg twice daily | Moderate | Repeated remote ischemic postconditioning (rIPostC) did not significantly reduce infarct size or improve clinical outcomes in ischemic stroke patients |
| 25 | Hougard et. Al. 2013 | Denmark | 443 | 285 | 285 | 66 | 4 | RIC | 247 | Sham | 196 | AIS | 1 | 4 | 5 | 18 | 22 | 4 inflations of an upper limb blood pressure cuff (200 mmHg) for 5 minutes, with 5 minutes of deflation between inflations. | Moderate | Remote ischemic perconditioning (rPerC) did not significantly improve infarct size or clinical outcomes but showed potential in tissue survival, especially in non-recanalized tissue. |
| 26 | Chen et. Al. 2022 | China | 1893 | 1170 | 723 | 65 | 3 | RIC | 922 | Sham | 971 | AIS | 14 |  |  | 48 | 48 | 5 cycles of 5 minutes ischemic and 5 minutes reperfusion to bilateral upper limbs, twice daily | Moderate | Treatment with remote ischemic conditioning significantly increased the likelihood of excellent neurologic function at 90 days in patients with acute moderate ischemic stroke |
| 27 | He et. Al. 2020 | China | 49 | 38 | 11 | 60 | 3 | RIC | 25 | Sham | 25 | AIS | 24 | 7 | 9 | 6 | 4 | Healthy upper limb; 4×(5 min ischemia at 200 mmHg + 5 min reperfusion); two sessions at ~6 h and ~18 h post-IVT; device BB-RIC-D1 (LAPUL). Sham: 60 mmHg | High | Adding two brief RIC sessions within 24 h after IVT in AIS was **safe**, did **not** improve clinical scales (mRS/NIHSS), but **reduced hs-CRP** at 24 h versus sham. |
| 28 | England et. Al. 2019 | United Kingdom | 60 | 36 | 24 | 72 | 3 | RIC | 31 | Sham | 29 | AIS | 1 | 7 | 7 | 254 | 195 | 4 cycles of limb ischemia (5 minutes of inflation, 5 minutes of deflation) using a BP cuff on the nonparetic arm. | Moderate | RIC is feasible and safe for use in hyperacute ischemic stroke, with potential clinical benefits requiring further evaluation in larger trials. |
| 29 | England et. Al. 2017 | United Kingdom | 26 | 17 | 9 | 76.2 | 3 | RIC | 13 | Sham | 13 | AIS | 15.8 | 6 | 6 | 15 | 15 | 4 cycles of intermittent limb ischemia, 5 minutes of inflation and 5 minutes of deflation performed manually using a blood pressure cuff on the non-paretic arm. | High | RIC is well-tolerated after acute stroke and may improve neurological outcomes. |
| 30 | Che et. Al. 2019 | China | 30 | 24 | 6 | 65.5 | 3 | RIC | 15 | Sham | 15 | AIS | 1 | 7 | 5 | 240 | 240 | Bilateral upper limbs, 5 cycles of 200 mmHg pressure inflation and deflation for 5 minutes, twice daily for 7 days. | Moderate | Remote ischemic postconditioning (RIPC) is feasible and safe for acute ischemic stroke (AIS) patients after intravenous rt-PA thrombolysis. |
| 31 | An et. al. 2024 | China | 23 | 18 | 5 | 54 | 3 | RIC | 13 | Sham | 10 | AIS | 3 |  |  | 120 | 120 | Bilateral upper limb ischemia for 5 cycles of 5 minutes each, with a 5-minute reperfusion period, twice a day for 3 months | Moderate | Remote ischemic conditioning (RIC) significantly improves cerebral hemodynamics in symptomatic intracranial atherosclerosis (sICAS) patients |
| 32 | Blauenfeldt et. Al. 2023 | Denmark | 902 | 567 | 335 | 72 | 3 | RIC | 436 | Sham | 466 | AIS | 7 | 6 | 5 | 6 | 6 | **Bilateral upper limb, 5 cycles, 50 minutes, 2 times per day** | High | RIC treatment did not significantly improve functional outcomes at 90 days in patients with acute stroke |
| 33 | Liang et. Al. 2023 | China | 132 | 71 | 35 | 64.5 | 3 | RIC | 57 | Sham | 49 | AIS | 30 | 4 | 4 | 18 | 18 | 4 cycles of 5-min inflation to 200 mmHg, 5 min deflation, once daily | High | RIPostC improved neurological outcomes in AIS patients, and autonomic function mediated this effect |
| 34 | Kerstens et. Al. 2023 | Netherlands | 88 | 60 | 28 | 72.8 | 3 | RIC | 40 | Sham | 48 | AIS | 14.5 | 5 | 6.5 | 14 | 18 | 4 cycles of 5-minute inflation followed by 5-minute deflation of the blood pressure cuff, twice daily | High | No significant difference was observed between repeated remote ischemic postconditioning (rIPostC) and sham-conditioning on quality of life, patient-reported outcomes, or clinical events in patients with ischemic stroke. |

Risk of Bias

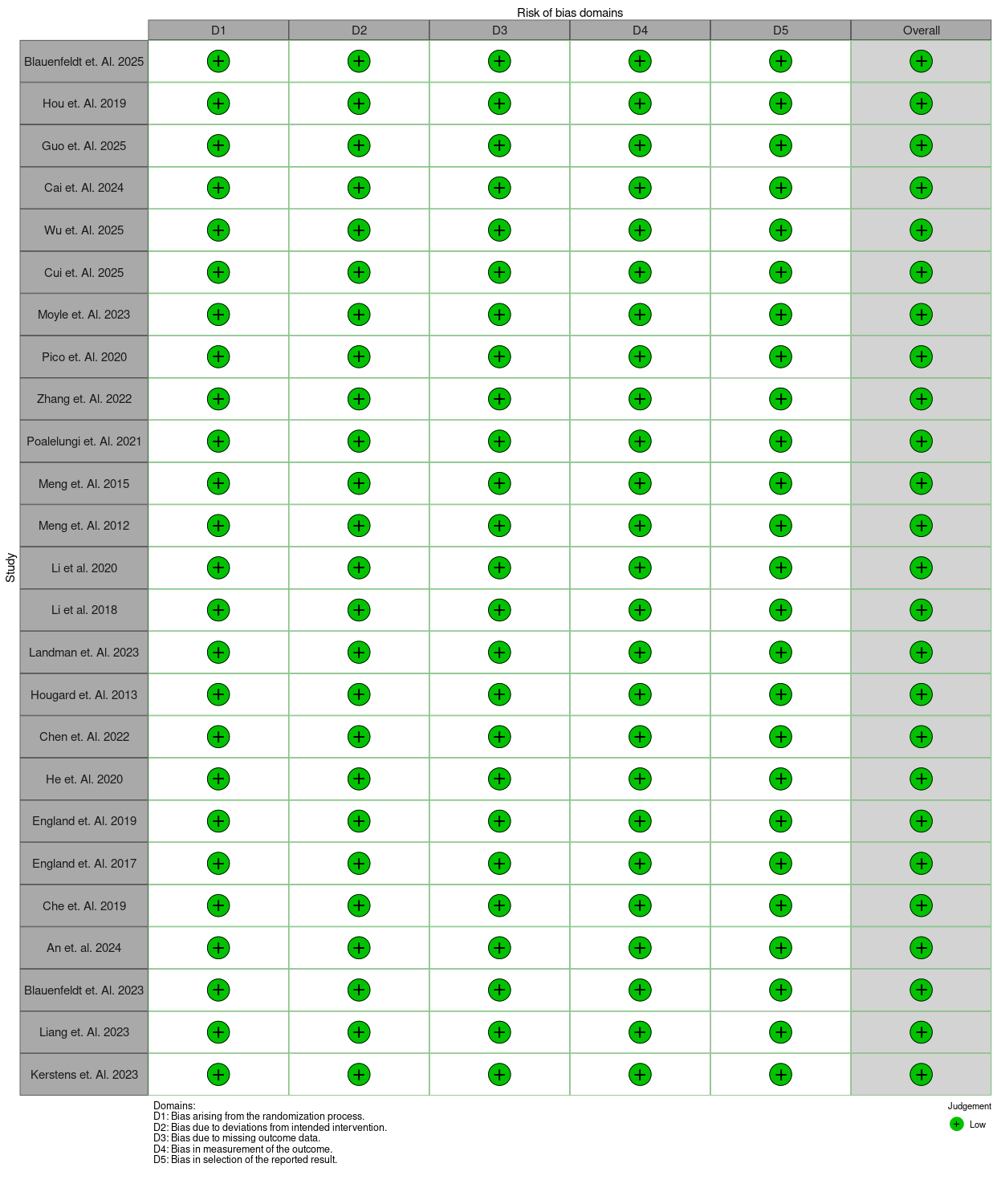
